# Image Based Deep Learning in 12-Lead ECG Diagnosis

**DOI:** 10.1101/2022.11.21.22282586

**Authors:** Raymond Ao, George He

## Abstract

**Background:** Most studies on machine learning classification of electrocardiogram (ECG) diagnoses focus on processing raw signal data rather than ECG images. In clinical practice, it is often the case where ECGs printed on paper or only digital images are easily accessible. This study aims to evaluate the accuracy of image based deep learning algorithms on 12 lead ECG diagnosis.

**Methods:** Deep learning models were trained on various 12-lead ECG datasets and evaluated for accuracy by testing on holdout test data as well as data from datasets not seen in training.

**Results:** The results demonstrated excellent AUROC, AUPRC, sensitivity and specificity on holdout test data from datasets used in training, but poorer accuracy on unseen datasets.

**Discussion:** This study demonstrates feasibility of image based deep learning algorithms in ECG diagnosis, and identifies directions for future research in order to develop clinically applicable deep-learning models in ECG diagnosis.

## 1 Introduction

Deep learning methods have been shown excellent diagnostic performance on classifying electrocardiogram (ECG) diagnoses using signal data, even surpassing individual cardiologist performance in some studies. Most studies published to date focus on machine learning diagnosis of ECGs by processing raw signal data. In clinical practice, obtaining signal data from ECG machines may be difficult, such as in cases where ECGs are printed on paper, or where only digital images are easily accessible. In this experiment, we trained convolutional neural networks (CNNs) capable of classifying raw images of 12-lead ECGs with comparable or superior performance to existing raw signal classifiers.

### 1.1 Literature review

Multiple studies have shown excellent diagnostic accuracy, including performance superior to cardiologists, in 12-lead ECG diagnosis. For example, one study which used raw ECG data created a DNN which performed similarly to or better than the average of individual cardiologists [1] in classifying 12 different rhythms, including atrial fibrillation/flutter, AVB, junctional rhythm, ventricular tachycardia, SVT, and Wenkebach in single lead ECG. Other studies used signal data from 12-lead ECGs with excellent results in arrhythmia classification.

The method used in this experiment differs from most other studies in that ECG image data is directly used to train and test deep learning models as opposed raw signal data or transformations of signal data. The benefit of this method is that models can be used in cases where there is no raw signal data (e.g. ECGs printed on paper) or where signal data is difficult to obtain and analyse. The only studies we have identified which use raw ECG images only differentiated between ‘normal’ and ‘sick’ or abnormal [2], or individual beats only [3]

## 2 Methods

### 2.1 Datasets

The primary dataset used for model development and evaluation was PTB-XL. We also tested for external validity of models across different unseen datasets, as well as with different datasets in combination.

The following publicly available ECG datasets were used:

- PTB-XL [4] (PTB)
- CPSC 2018 database [5] (CPSC)
- 12-lead ECG database for arrhythmia research from Chapman University and Shaoxing People’s Hospital [6] (Shaoxing)
- Test dataset for: Automatic multi-label ECG diagnosis of impulse or conduction abnormalities in patients with deep learning algorithm: a cohort study [7] (Tongji)

Each dataset contained raw 10-second 12-lead ECG signal waveform data with corresponding diagnostic labels.

### 2.2 Data pre-processing

For each individual ECG sample, an image was generated by plotting the signal data. We used the Python ECG plot [8] library, which generates 12-lead ECG images resembling ECG displays and print-outs commonly used in clinical practice. An example is shown in Figure 1. Images were processed at 1600x512 resolution.

**Figure 1:**
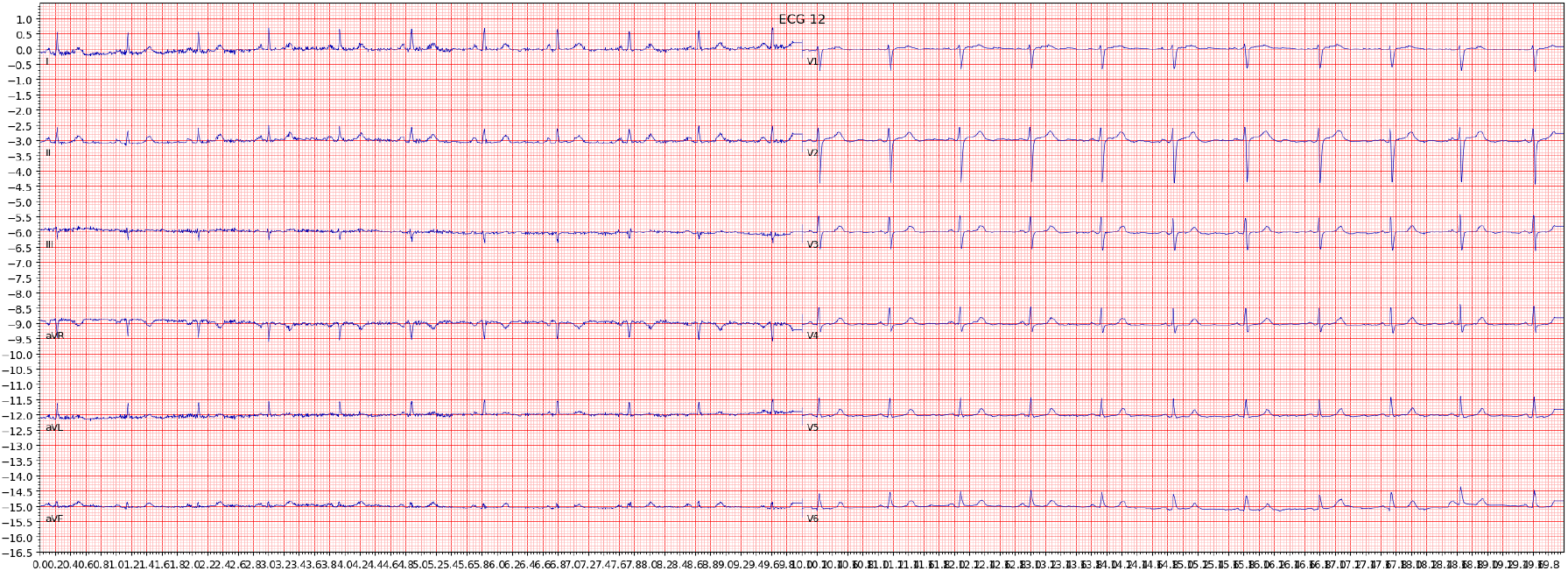
Example of an ECG image generated from the signal data of one sample in the dataset

The images were then converted to grayscale, then binarised using simple thresholding, Otsu thresholding, and adaptive thresholding. A binarised copy was saved for each of the thresholding techniques. For further data augmentation, a slightly blurred vision of the grayscale image was created, and the aforementioned thresholding techniques were also applied. In total, eight augmented copies of each original ECG image was generated. An example of an image after grayscale conversion and adaptive thresholding is shown in figure 2.

**Figure 2:**
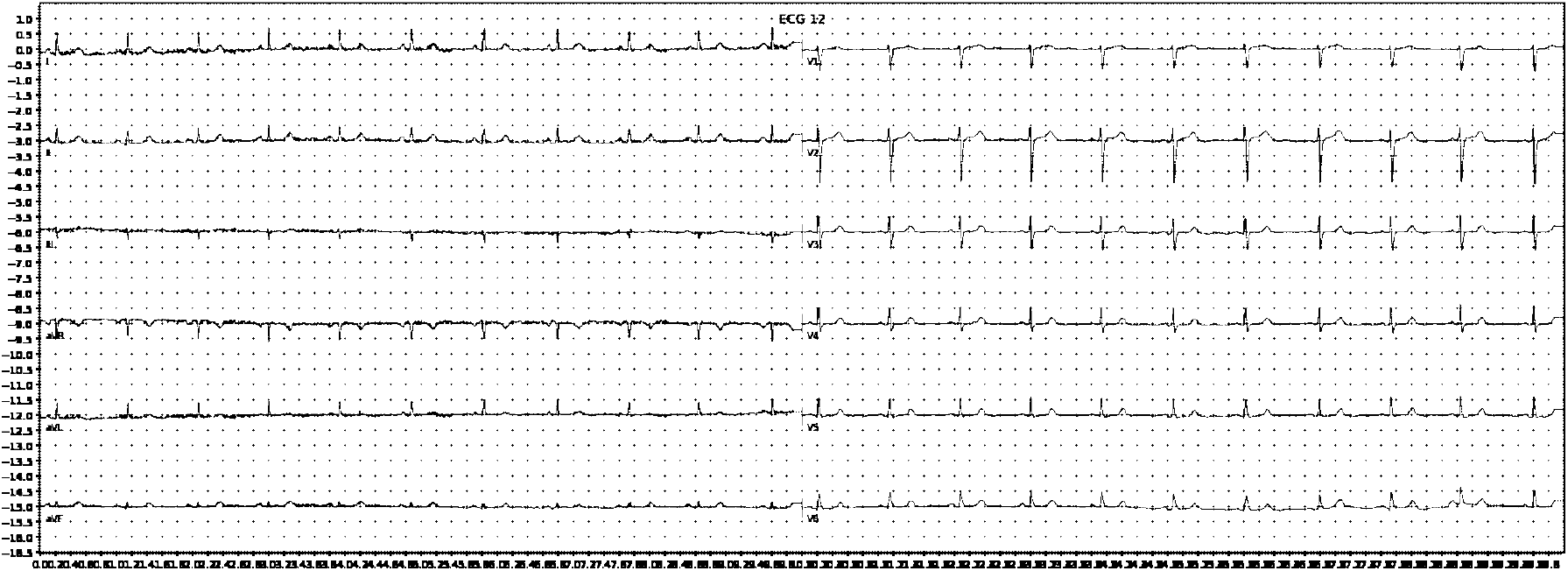
Example of an ECG image after grayscale conversion and adaptive thresholding

**Figure 3:**
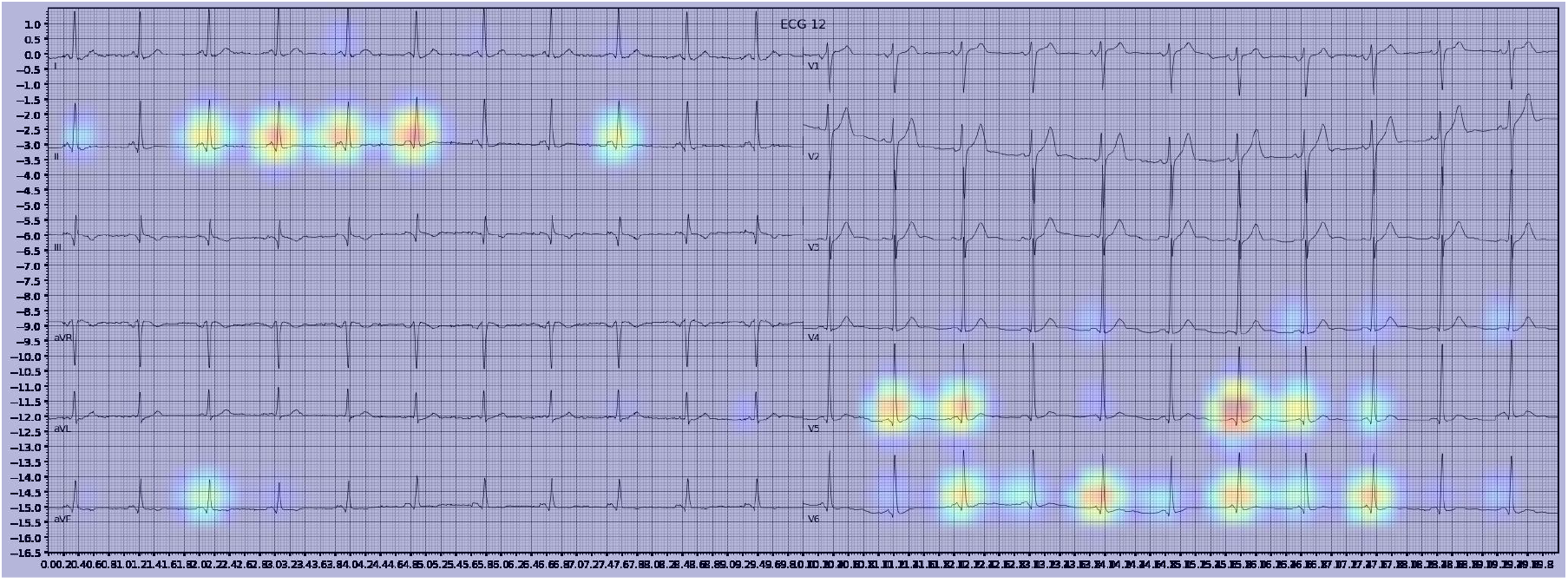
Example of an activation heatmap for an ECG showing WPW

**Figure 4:**
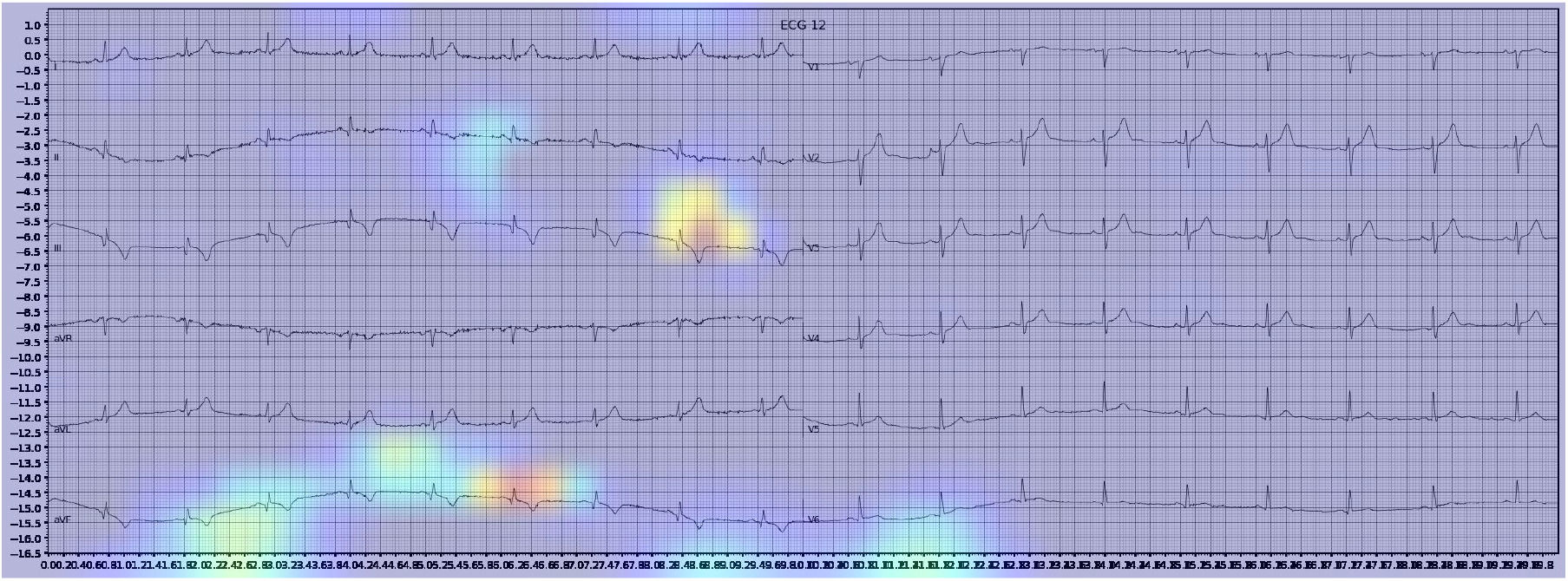
Example of an activation heatmap for an ECG showing myocardial infarction

**Figure 5:**
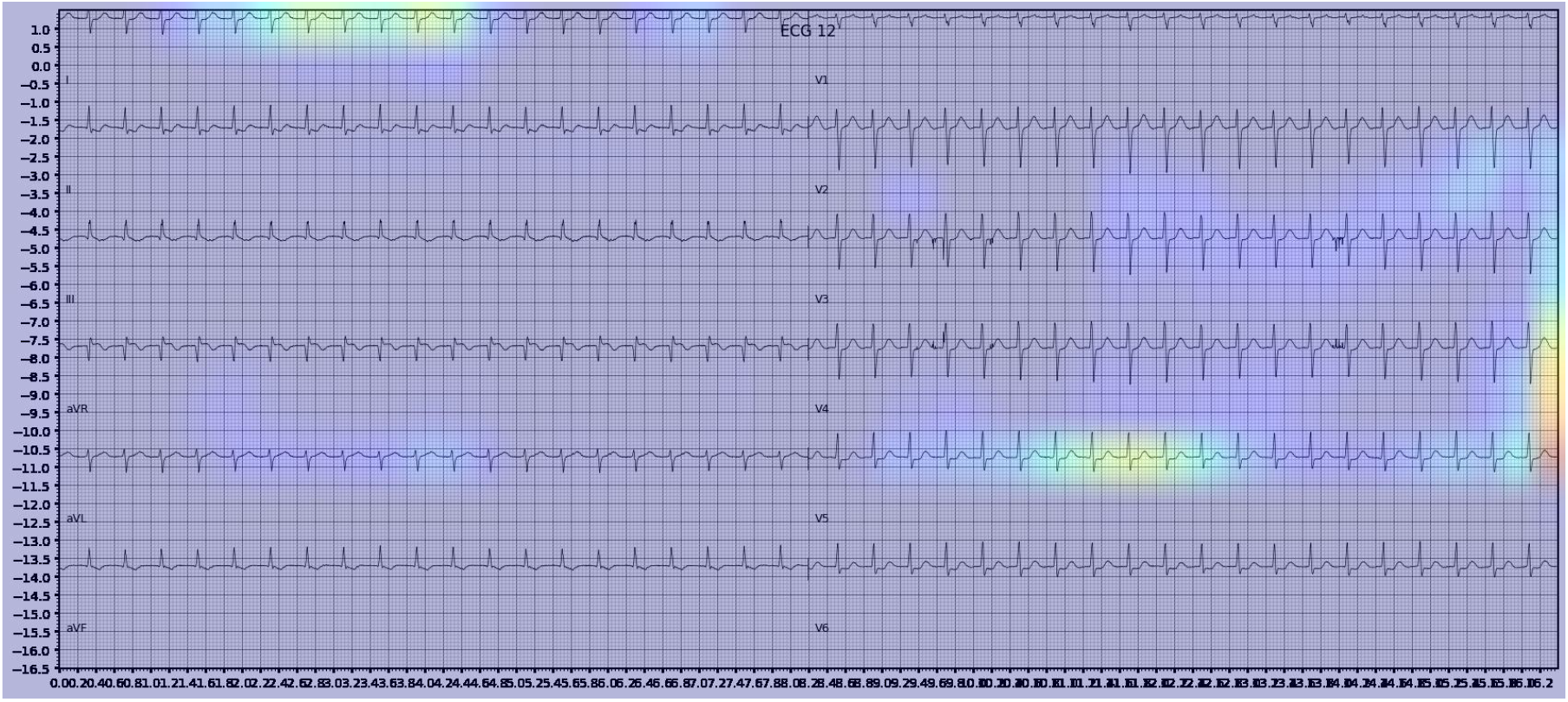
Example of an activation heatmap for an ECG showing SVT

### 2.3 Classification task overview

Binary classification models were trained predict the presence and absence of:

- Normal ECG (NORM)
- Left bundle branch block (LBBB)
- Right bundle branch block (RBBB)
- Atrial fibrillation (AFIB)
- Atrial flutter (AFLT)
- First degree AV block (fAVB)
- Myocardial infarction (MI)
- Wolff-Parkinson White (WPW)
- Supraventricular tachycardia

Models were trained using the PTB-XL dataset and evaluated on holdout test data from PTB-XL. Additionally, models were also tested on ECG images from other datasets not involved in training. Further testing was done on combined datasets, where matching diagnostic labels were present.

For each training run, the included samples from all datasets were randomly shuffled, and split into training, validation and holdout test sets, with splits of 0.8, 0.1 and 0.1 respectively.

### 2.4 Model architecture and training

Model were built on VGG16 architecture with Imagenet pre-trained weights. The original classification layer was removed and replaced with a classification head consisting of a global average pooling 2D layer, a dropout layer for training, followed by a fully connected layer with one output and sigmoid activation.

The classification head was initially trained for up to 10 epochs with early stopping, while all other layers were frozen. The entire model was then unfrozen, and trained until no further drop in validation loss was seen (early stopping with patience of 6). The learning rate was 1x 10-^5. A learning rate schedule involving reducing the learning rate when the validation loss plateaued was trialled, without significant improvement of results.

For most training instances, binary cross-entropy loss was used. We also experimented with focal loss for highly imbalanced datasets.

## 3 Results

The models demonstrated good performance when tested on unseen holdout test data from the original datasets used on training. Generalisation to unseen, external datasets was poorer. Performance of models trained on a combination of different datasets mixed together showed good performance on holdout test splits containing the mixed datasets. The results are summarised in table below

**Table 1:** Test results of models trained on PTB-XL ECGs and tested on a holdout test set from PTB-XL

| Diagnosis | AUPRC | AUROC | Sensitivity | Specificity |
| --- | --- | --- | --- | --- |
| AFIB | 0.999 | 1.000 | 0.992 | 0.999 |
| AFLT | 0.985 | 0.989 | 1.000 | 0.964 |
| AMI | 0.9895 | 0.991 | 0.977 | 0.944 |
| AVB | 0.979 | 0.985 | 0.969 | 0.940 |
| IMI | 0.981 | 0.983 | 0.948 | 0.9162 |
| LMI | 0.540 | 0.511 | 0.349 | 0.632 |
| LVH | 0.934 | 0.971 | 0.897 | 0.954 |
| MI | 0.988 | 0.989 | 0.960 | 0.940 |
| NORM | 0.997 | 0.998 | 0.980 | 0.980 |
| PVC | 0.984 | 0.986 | 0.950 | 0.970 |
| RBBB | 1.000 | 1.000 | 0.996 | 1.000 |
| sAVB | 0.860 | 0.960 | 0.800 | 0.933 |
| Sex (Female) | 0.997 | 0.996 | 0.985 | 0.954 |
| SVT | 1.000 | 1.000 | 0.875 | 1.000 |
| tAVB | 0.848 | 0.946 | 1.000 | 0.867 |

**Table 2:** Test results of models trained on combined datasets and tested on holdout data from the combined datasets

| Diagnosis | Dataset combination | AUPRC | AUROC | Sensitivity | Specificity |
| --- | --- | --- | --- | --- | --- |
| AFIB | PTB+CPSC+Shaoxing | 0.994 | 0.999 | 0.970 | 1.00 |
| fAVB | PTB+CPSC+Shaoxing | 0.985 | 0.987 | 0.95 | 0.960 |
| LBBB | PTB+CPSC+Shaoxing | 0.987 | 0.999 | 0.940 | 1.000 |
| RBBB | PTB+CPSC+Shaoxing | 0.298 | 0.739 | 0.460 | 0.850 |
| MI | PTB+Shaoxing | 0.954 | 0.987 | 0.800 | 0.990 |
| SVT | PTB+Shaoxing | 0.980 | 1.000 | 0.950 | 1.000 |
| WPW | PTB+Shaoxing+Tongji | 0.756 | 0.842 | 0.930 | 0.370 |

**Table 3:**
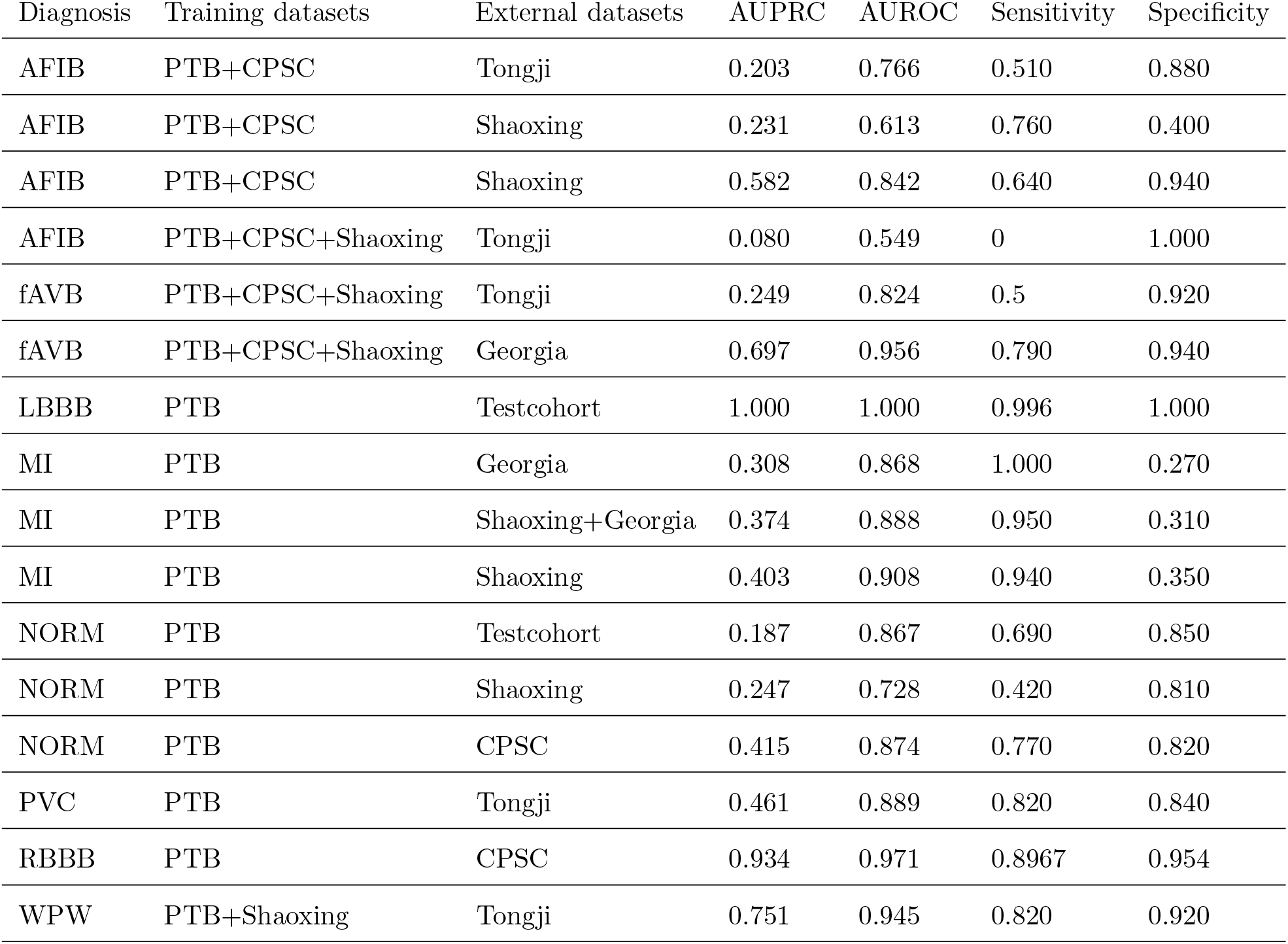
Test results of models tested on separate, unseen datasets than those used in training

| Diagnosis | Training datasets | External datasets | AUPRC | AUROC | Sensitivity | Specificity |
| --- | --- | --- | --- | --- | --- | --- |
| AFIB | PTB+CPSC | Tongji | 0.203 | 0.766 | 0.510 | 0.880 |
| AFIB | PTB+CPSC | Shaoxing | 0.231 | 0.613 | 0.760 | 0.400 |
| AFIB | PTB+CPSC | Shaoxing | 0.582 | 0.842 | 0.640 | 0.940 |
| AFIB | PTB+CPSC+Shaoxing | Tongji | 0.080 | 0.549 | 0 | 1.000 |
| fAVB | PTB+CPSC+Shaoxing | Tongji | 0.249 | 0.824 | 0.5 | 0.920 |
| fAVB | PTB+CPSC+Shaoxing | Georgia | 0.697 | 0.956 | 0.790 | 0.940 |
| LBBB | PTB | Testcohort | 1.000 | 1.000 | 0.996 | 1.000 |
| MI | PTB | Georgia | 0.308 | 0.868 | 1.000 | 0.270 |
| MI | PTB | Shaoxing+Georgia | 0.374 | 0.888 | 0.950 | 0.310 |
| MI | PTB | Shaoxing | 0.403 | 0.908 | 0.940 | 0.350 |
| NORM | PTB | Testcohort | 0.187 | 0.867 | 0.690 | 0.850 |
| NORM | PTB | Shaoxing | 0.247 | 0.728 | 0.420 | 0.810 |
| NORM | PTB | CPSC | 0.415 | 0.874 | 0.770 | 0.820 |
| PVC | PTB | Tongji | 0.461 | 0.889 | 0.820 | 0.840 |
| RBBB | PTB | CPSC | 0.934 | 0.971 | 0.8967 | 0.954 |
| WPW | PTB+Shaoxing | Tongji | 0.751 | 0.945 | 0.820 | 0.920 |

## 4 Visual explanation

Gradient-weighted Class Activation Mapping (Grad-CAM) creates a heatmap to visualise areas of the image which are important in predicting its class.

Here are examples of activation maps

## 5 Discussion

The results show that computer vision models are capable of achieving good diagnostic performance on unseen ECGs sampled from the same population(s) and dataset(s) as that used for model training, including combinations of different datasets. The models were not able to generalise well to unseen external datasets - this may be due to differences in labelling criteria for diagnoses between the datasets, or variances in ECG quality. Overall, classification performance on ECG images using deep CNNs is comparable to the best models using raw ECG signal holdout test data from the same dataset.

### 5.1 Future work and application

This research demonstrates that computer vision AI models can diagnose conditions on ECG with good accuracy. Future research which could bring this technology closer to clinical application could focus on developing models which can generalise to a wide range of ECG image formats from various sources, and cover a wider range of relevant clinical diagnoses. Additional diagnoses which would be of interest clinically would include diagnosing STEMI in patients with LBBB or pacemaker, differentiating SVT with abberency vs VT, and specific subtypes of AV block. Furthermore, models could be developed to be applicable to different ECG formats or styles. The techniques demonstrated here could also be applied for novel practical applications, such as smartphone applications to diagnose photos of ECGs, or in telehealth.

## Data Availability

Data used is publicly available

http://2018.icbeb.org/Challenge.html

https://physionet.org/content/ptb-xl/1.0.3/

https://pubmed.ncbi.nlm.nih.gov/32051412/

